# Foundation Model Robustness to Technical Acquisition Parameters in Chest X-Ray AI: A Multi-Architecture Comparative Study with External Validation

**DOI:** 10.64898/2026.01.25.26344809

**Authors:** Hayden Farquhar

## Abstract

**Background:** Foundation models have emerged as a promising paradigm for medical imaging AI [7], with claims of improved generalization and reduced bias. However, their robustness to technical acquisition parameters remains unexplored. We evaluated whether foundation models exhibit greater robustness to chest radiograph view type (anteroposterior [AP] versus posteroanterior [PA]) compared to traditional convolutional neural networks.

**Methods:** We compared four model architectures on the RSNA Pneumonia Detection Challenge dataset (n=26,684 images) and externally validated on the NIH ChestX-ray14 dataset (n=112,120 images): DenseNet-121 (supervised CNN), BiomedCLIP (vision-language model trained on 15 million biomedical image-text pairs), RAD-DINO (self-supervised model trained on 5+ million radiographs), and CheXzero (vision-language model trained on MIMIC-CXR chest radiographs). Primary outcome was the sensitivity gap between AP and PA views, with bootstrap confidence intervals and permutation testing.

**Results:** On RSNA, CheXzero showed the smallest gap (14.3%, 95% CI: 11.2-17.5%), followed by RAD-DINO (25.2%, 22.6-27.9%), DenseNet-121 (35.7%, 32.9-38.7%), and BiomedCLIP (36.1%, 33.5-39.0%). However, on external validation (NIH), **model rankings reversed completely**: RAD-DINO demonstrated the smallest gap (22.3%, 95% CI: 21.0-23.6%), while CheXzero’s gap **increased dramatically to 48.9%** (95% CI: 47.7-50.1%). Domain-specific training provided robustness within the training domain but failed to generalize. On PA view pneumonia cases in NIH, 31% were missed by all four models, representing a systematic blind spot. View type explained 61-100% of performance variance across models on both datasets, compared to 0-38% for age and less than 4% for sex.

**Conclusions:** Foundation models do not eliminate technical acquisition parameter biases in chest X-ray AI. While domain-specific training (CheXzero) provided superior robustness on internal validation, this advantage **collapsed on external data**. Self-supervised learning (RAD-DINO) demonstrated the most generalizable robustness, with consistent view type gap stability across datasets with different labeling schemes (25.2% → 22.3%, despite substantial AUC differences). These findings challenge assumptions about foundation model generalization and highlight the need for acquisition parameter auditing in AI regulatory frameworks and multi-site external validation for robustness claims.

## Introduction

Artificial intelligence systems for chest radiograph interpretation are increasingly deployed in clinical practice [1], with applications ranging from pneumonia detection to tuberculosis screening. The landmark CheXNet study demonstrated that a 121-layer DenseNet could achieve radiologist-level pneumonia detection on the NIH ChestX-ray14 dataset, catalyzing rapid development of deep learning approaches for chest radiograph analysis. However, concerns about algorithmic fairness [12] have focused predominantly on demographic factors such as age, sex, and race, while technical acquisition parameters have received comparatively little attention.

Chest radiographs can be acquired in anteroposterior (AP) or posteroanterior (PA) projection [14], with important implications for image characteristics. PA views, acquired with the patient standing and the detector against their chest, produce sharper cardiac borders and less magnification. AP views, typically acquired at the bedside in patients too ill to stand, show cardiac magnification and may have lower image quality. Critically, AP imaging is disproportionately used in sicker patients, creating a confounded relationship between acquisition method and disease severity.

Foundation models have emerged as a promising paradigm for medical imaging AI [7]. These models, pre-trained on large datasets using self-supervised or vision-language learning, are claimed to offer improved generalization across diverse populations and imaging conditions. BiomedCLIP, trained on 15 million biomedical image-text pairs from PubMed Central articles, represents the general biomedical foundation model approach. CheXzero demonstrated that contrastive learning on MIMIC-CXR image-text pairs enables zero-shot multi-label pathology classification at radiologist-level accuracy without explicit annotations. RAD-DINO employs self-supervised learning on over 800,000 chest radiographs without text supervision.

Despite this growing literature on demographic bias [2,3,8,9], no study has systematically examined foundation model robustness to technical acquisition parameters. This represents a critical gap given the documented impact of acquisition factors on traditional CNN performance and the dominance of technical over demographic factors in determining AI performance variance. We previously demonstrated that view type explains 69-87% of performance variance in supervised CNN models for pneumonia detection, far exceeding the contribution of demographic factors. This study extends that work to foundation models, asking: Do modern foundation models exhibit greater robustness to acquisition parameter variation than traditional architectures? And critically, does any observed robustness advantage generalize to external datasets?

## Methods

### Study Design and Datasets

We conducted a retrospective comparative study using two publicly available chest radiograph datasets with expert annotations and view position metadata.

Primary Dataset (RSNA): The RSNA Pneumonia Detection Challenge dataset provides expert annotations of possible pneumonia on chest radiographs [10] from the NIH Clinical Center. The dataset contains 26,684 chest radiographs with bounding box annotations for pneumonia-positive cases from 18 board-certified radiologists and metadata including view position (AP or PA), patient age, and patient sex extracted from DICOM headers. The dataset included 12,173 AP views (45.7%) and 14,511 PA views (54.3%), with an overall pneumonia prevalence of 22.5%. Pneumonia prevalence differed substantially by view type: 38.3% in AP views versus 9.3% in PA views, consistent with AP imaging being preferentially used in sicker patients.

External Validation Dataset (NIH): The NIH ChestX-ray14 dataset [15] contains 112,120 frontal chest radiographs from 30,805 unique patients with 14 disease labels extracted from radiology reports using natural language processing. View position metadata is available in the dataset. The dataset included 68,418 AP views (61.0%) and 43,702 PA views (39.0%). For pneumonia detection, we used “Infiltration” as the primary proxy label (prevalence: 17.7%), consistent with prior literature, with sensitivity analyses using “Consolidation,” explicit “Pneumonia” labels, and combined definitions.

### Model Selection

We evaluated four model architectures representing distinct training paradigms:

DenseNet-121 (Supervised CNN): A convolutional neural network trained with supervised learning on multiple chest X-ray datasets using the TorchXRayVision library [16].

BiomedCLIP [4] (Vision-Language, General Biomedical): A CLIP-style vision-language model trained on PMC-15M, a dataset of 15 million biomedical image-text pairs from PubMed Central articles spanning diverse imaging modalities.

RAD-DINO [6] (Self-Supervised, Radiology-Specific): A vision transformer pretrained on 882,775 chest X-rays using DINOv2 self-supervised learning without text supervision.

CheXzero [5] (Vision-Language, Chest X-Ray Specific): A CLIP-style vision-language model trained on 377,110 MIMIC-CXR chest radiographs paired with radiology reports.

### Model Evaluation

Vision-language models (BiomedCLIP, CheXzero) were evaluated using zero-shot classification with text prompts, following the contrastive learning paradigm established by CLIP. For RAD-DINO, which lacks text encoding capability, we trained a linear probe on extracted embeddings using logistic regression. DenseNet-121 was evaluated using its native pneumonia prediction output.

### Statistical Analysis

The primary outcome was the sensitivity gap between AP and PA views, defined as Sensitivity(AP) minus Sensitivity(PA). Optimal classification thresholds were determined using Youden’s J statistic. Bootstrap resampling (n=2,000 iterations) generated 95% confidence intervals using stratified sampling to preserve view type and disease status distribution. Secondary outcomes included AUC, calibration metrics (expected calibration error [ECE], Brier score), variance decomposition analysis, and inter-model agreement analysis. For external validation comparison, we assessed whether 95% confidence intervals overlapped between datasets to determine statistical significance of performance changes.

## Results

### Primary Analysis (RSNA Dataset)

All four models demonstrated statistically significant performance gaps between AP and PA views on the RSNA dataset (all p<0.001, Table 1). CheXzero exhibited the smallest gap at 14.3% (95% CI: 11.2-17.5%), followed by RAD-DINO at 25.2% (22.6-27.9%), DenseNet-121 at 35.7% (32.9-38.7%), and BiomedCLIP at 36.1% (33.5-39.0%). Pairwise comparisons revealed significant differences between model architectures. CheXzero significantly outperformed all other models (all p<0.001), with sensitivity gap reductions of 10.9 percentage points versus RAD-DINO, 21.4 points versus DenseNet-121, and 21.9 points versus BiomedCLIP. The most striking finding was the differential performance between the two vision-language models: CheXzero, trained on chest X-ray-specific data, demonstrated a 2.5-fold smaller view type gap compared to BiomedCLIP, trained on general biomedical images, despite sharing an identical CLIP-based architecture.

**Table 1.** View Type Performance Across Model Architectures (RSNA Dataset)

| Model | AUC | AP Sens (%) | PA Sens (%) | Gap (95% CI) | P-value |
| --- | --- | --- | --- | --- | --- |
| CheXzero | 0.830 | 89.7 | 75.4 | 14.3 (11.2-17.5) | <0.001 |
| RAD-DINO | 0.905 | 90.9 | 65.7 | 25.2 (22.6-27.9) | <0.001 |
| DenseNet-121 | 0.813 | 80.6 | 44.9 | 35.7 (32.9-38.7) | <0.001 |
| BiomedCLIP | 0.848 | 90.3 | 54.2 | 36.1 (33.5-39.0) | <0.001 |

*Abbreviations: AUC, area under the receiver operating characteristic curve; AP, anteroposterior; PA, posteroanterior; Sens, sensitivity; CI, confidence interval.*

### External Validation (NIH ChestX-ray14)

External validation on the NIH ChestX-ray14 dataset (n=112,120) revealed a critical limitation of domain-specific training: **the performance advantage did not generalize** (Table 2, Figure 2). Model rankings reversed completely on external data. RAD-DINO demonstrated the smallest gap (22.3%, 95% CI: 21.0-23.6%), showing remarkable consistency with RSNA (change: −2.9 percentage points). In contrast, CheXzero’s gap **increased dramatically from 14.3% to 48.9%** (change: +34.6 percentage points, 95% CIs non-overlapping, p<0.001). BiomedCLIP showed moderate deterioration (36.1% → 49.5%, +13.4 points), while DenseNet-121 remained relatively stable (35.7% → 38.9%, +3.2 points).

**Table 2.** Cross-Dataset Comparison: RSNA vs NIH External Validation.

| Model | RSNA Gap (95% CI) | NIH Gap (95% CI) | $\Delta$ Gap | $p < 0.05?$ |
| --- | --- | --- | --- | --- |
| RAD-DINO | 25.2% [22.6-27.9] | 22.3% [21.0-23.6] | -2.9% | No |
| DenseNet-121 | 35.7% [32.9-38.7] | 38.9% [37.7-40.2] | +3.2% | No |
| CheXzero | 14.3% [11.2-17.5] | 48.9% [47.7-50.1] | +34.6% | Yes*** |
| BiomedCLIP | 36.1% [33.5-39.0] | 49.5% [48.3-50.7] | +13.4% | Yes*** |

\*\*\* = 95% CIs non-overlapping (significant difference between datasets). Models sorted by external validation performance.

**Table 3.** NIH External Validation: Complete Results.

| Model | AUC | AP Sens | PA Sens | Gap (95% CI) | ECE AP | ECE PA |
| --- | --- | --- | --- | --- | --- | --- |
| RAD-DINO | 0.652 | 73.5% | 51.2% | 22.3% [21.0-23.6] | 0.275 | 0.280 |
| DenseNet-121 | 0.680 | 72.5% | 33.6% | 38.9% [37.7-40.2] | 0.391 | 0.479 |
| CheXzero | 0.664 | 78.8% | 29.9% | 48.9% [47.7-50.1] | 0.441 | 0.262 |
| BiomedCLIP | 0.639 | 80.0% | 30.5% | 49.5% [48.3-50.7] | 0.326 | 0.211 |

NIH ChestX-ray14 dataset (n=112,120). Infiltration used as pneumonia proxy. Models sorted by view type gap.

**Figure 1.**
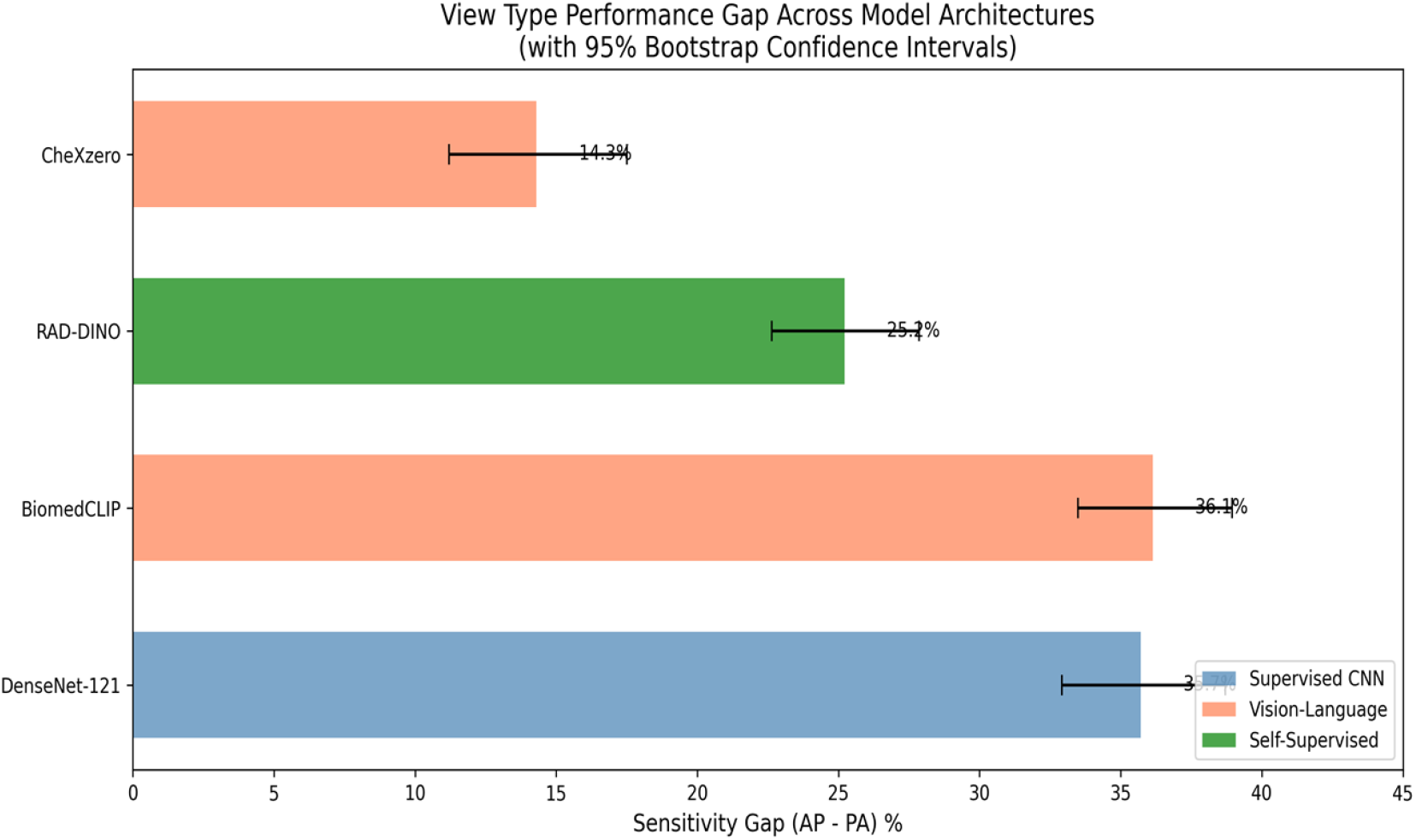
View Type Performance Gap Across Model Architectures (RSNA Dataset). Forest plot showing sensitivity gap (AP minus PA) with 95% bootstrap confidence intervals for each model. CheXzero demonstrates the smallest gap (14.3%), while BiomedCLIP and DenseNet-121 show the largest gaps (>35%).

**Figure 2.**
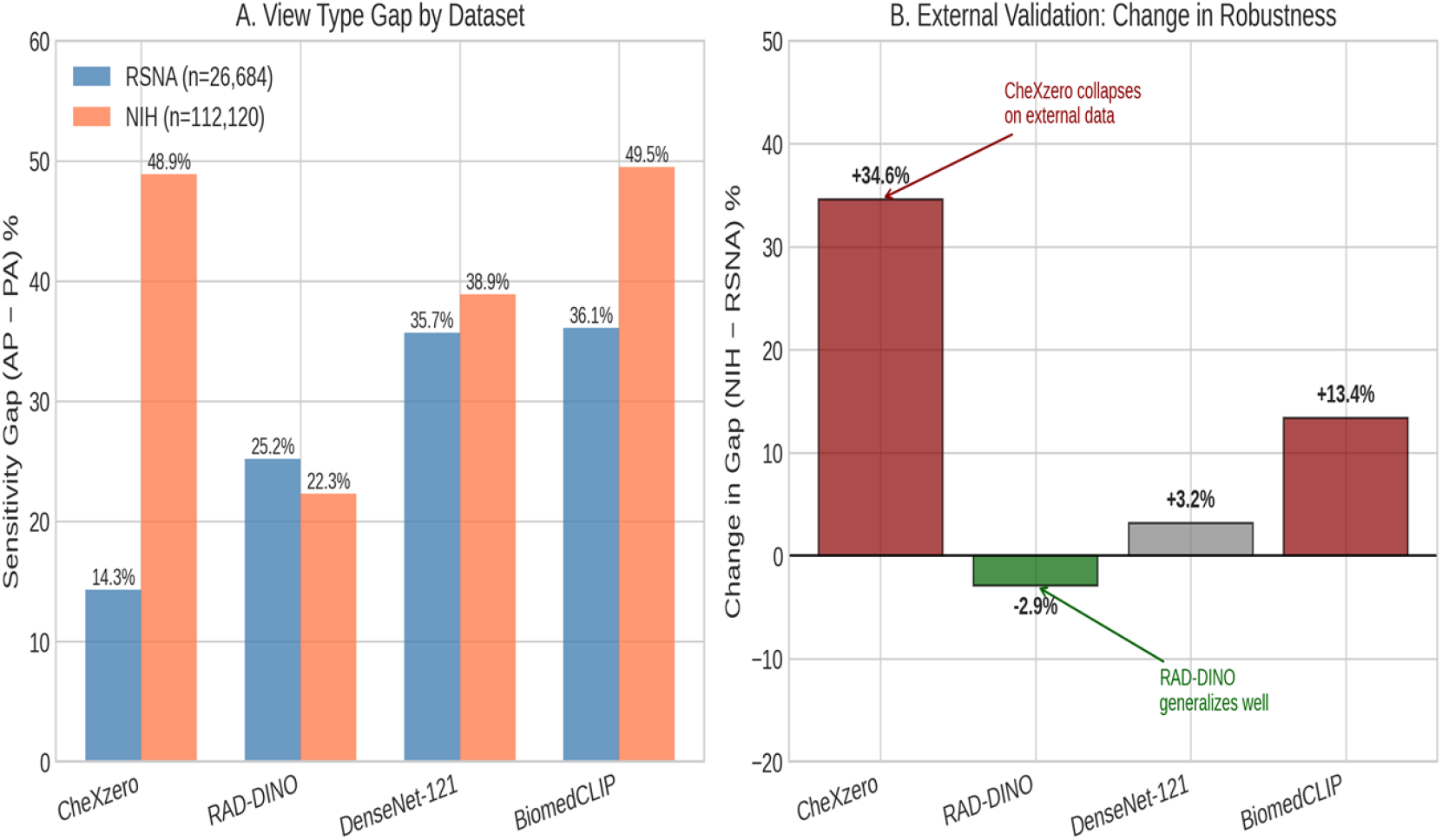
Cross-Dataset Comparison: RSNA vs NIH External Validation. (A) View type sensitivity gap on RSNA (blue) and NIH (orange) datasets for each model. (B) Change in gap between datasets, highlighting CheXzero’s dramatic deterioration (+34.6%) and RAD-DINO’s stability (−2.9%). CheXzero’s domain-specific advantage did not generalize to external data with different labeling conventions.

### Model Agreement Analysis

Analysis of inter-model agreement revealed a concerning finding: on PA view pneumonia cases in the NIH dataset, **31% were missed by all four models** compared to only 6% on AP views (Figure 3). This systematic blind spot suggests that PA view pneumonia detection represents a fundamental challenge across all architectures, not merely a model-specific limitation. Inter-model agreement (Cohen’s κ) was lower on PA views (κ=0.156-0.719) compared to AP views (κ=0.338-0.635), indicating greater disagreement in the challenging PA subset.

**Figure 3.**
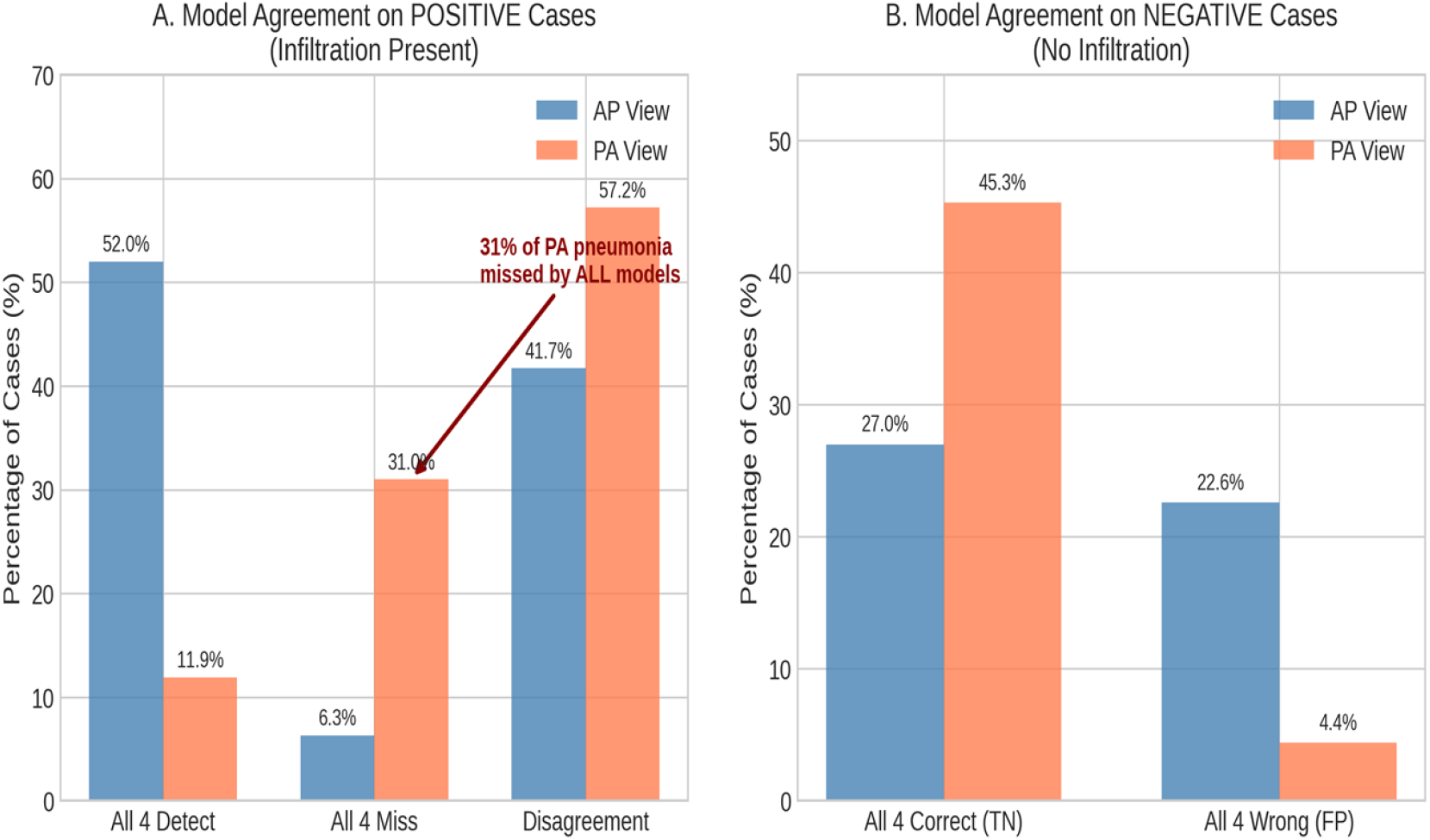
Inter-Model Agreement by View Type (NIH Dataset). (A) Positive cases: On PA views, 31% of pneumonia cases were missed by all four models compared to only 6% on AP views, indicating a systematic blind spot. (B) Negative cases: Higher specificity on PA views (45% all correct) compared to AP views (27% all correct).

### Alternative Label Definitions

To assess whether findings were specific to the “Infiltration” label used as a pneumonia proxy, we evaluated RAD-DINO across six pneumonia-related label definitions (Figure 4). The view type gap persisted across all definitions: Infiltration (22.3%), Consolidation (17.3%), explicit Pneumonia (24.5%), and various combinations (21.3-22.4%). This demonstrates that the view type bias is not an artifact of label choice but a fundamental challenge.

**Figure 4.**
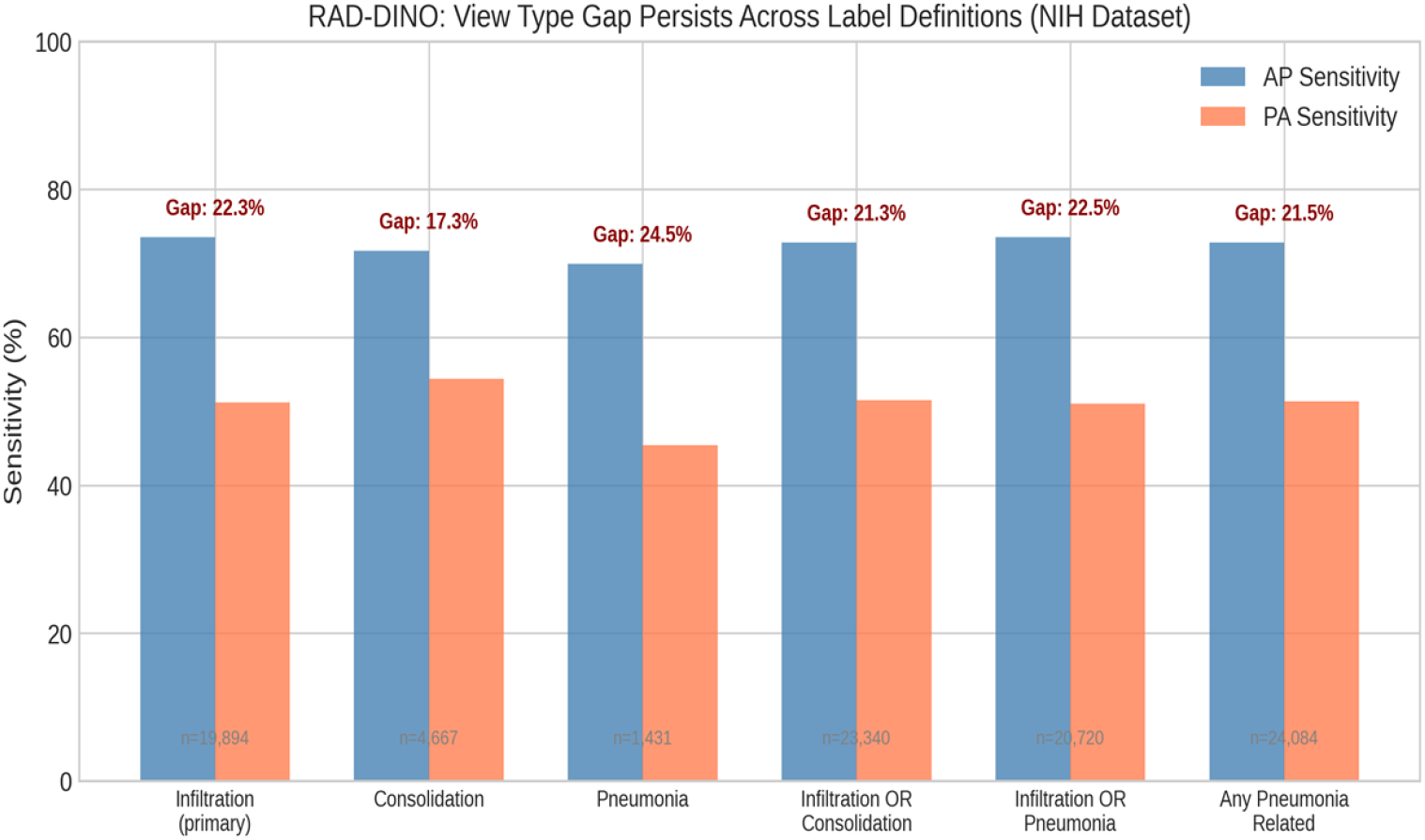
Alternative Label Definitions (RAD-DINO on NIH). Sensitivity by view type across six pneumonia-related label definitions in NIH ChestX-ray14. The view type gap (17-25%) persists regardless of label choice, demonstrating this is a fundamental challenge rather than a labeling artifact. Sample sizes shown for each definition.

### Variance Decomposition

Variance decomposition analysis confirmed that view type dominated performance variation across all models on both datasets (Table 4, Figure 5). On RSNA, view type explained 76-100% of variance. On NIH, view type explained 61-88% of variance. In contrast, age explained only 0-38% of variance, and sex contributed less than 4% across all models. These findings are consistent across datasets and model architectures.

**Table 4.**
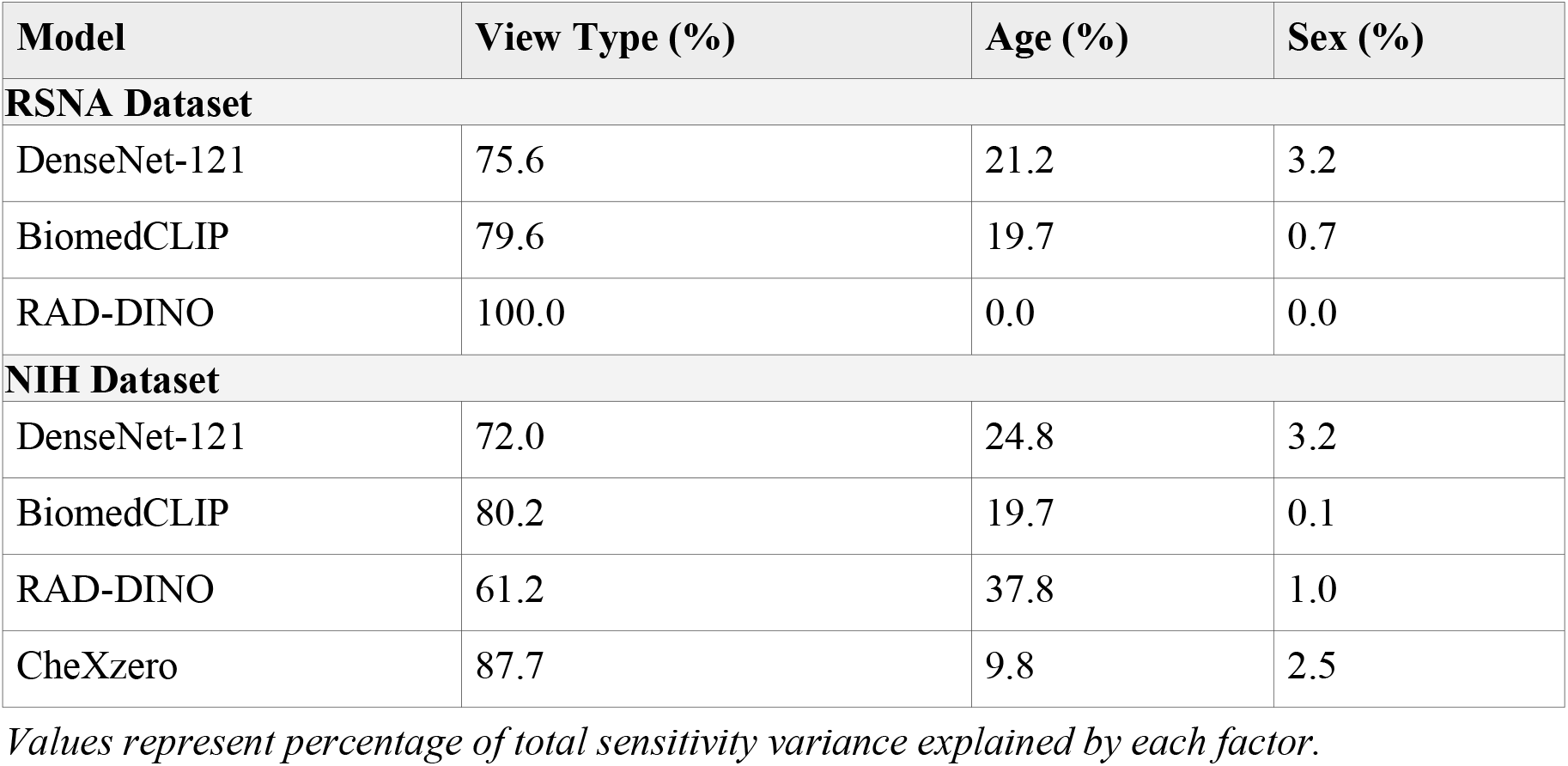
Variance Decomposition: Technical vs Demographic Factors.

**Figure 5.**
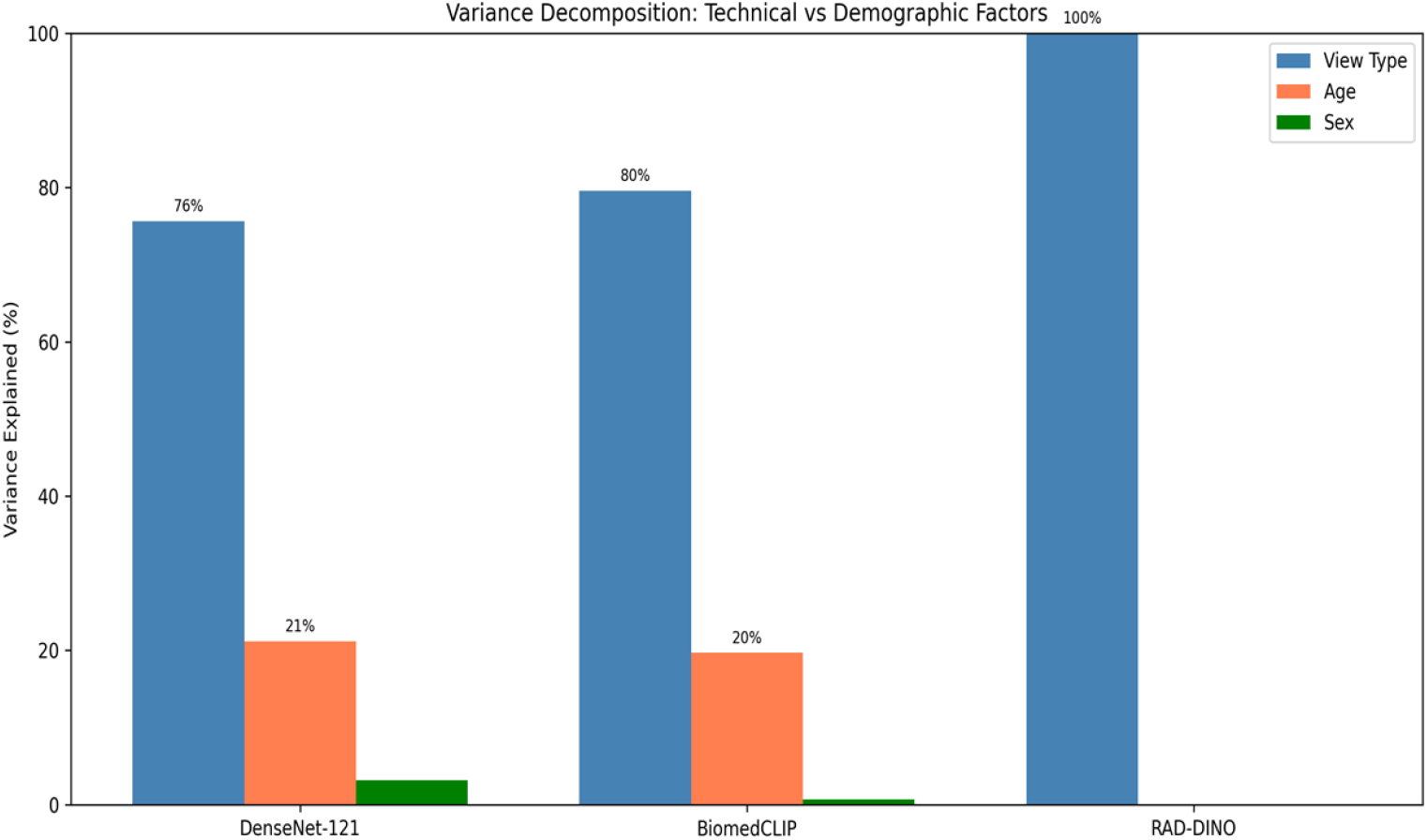
Variance Decomposition: Technical vs Demographic Factors. View type (blue) explains 76-100% of sensitivity variance on RSNA and 61-88% on NIH, while age (orange) contributes 0-38% and sex (green) less than 4%. This dominance of technical over demographic factors is consistent across architectures and datasets.

### Calibration Analysis

Model calibration varied substantially across architectures and view types (Figure 6). RAD-DINO demonstrated excellent calibration stability across view types (ECE difference: <0.01 on RSNA, 0.005 on NIH). In contrast, DenseNet-121 showed marked differential calibration, with substantially worse ECE on PA views (0.428 on RSNA, 0.479 on NIH) compared to AP views (0.218, 0.391). CheXzero and BiomedCLIP showed paradoxically better calibration on PA views than AP views on NIH data; this occurs because when models consistently output low probabilities for PA pneumonia cases, these probabilities may statistically align with the observed frequency of the majority class (negatives), yielding low ECE despite failing to assign sufficiently high probabilities to positive cases to cross decision thresholds.

**Figure 6.**
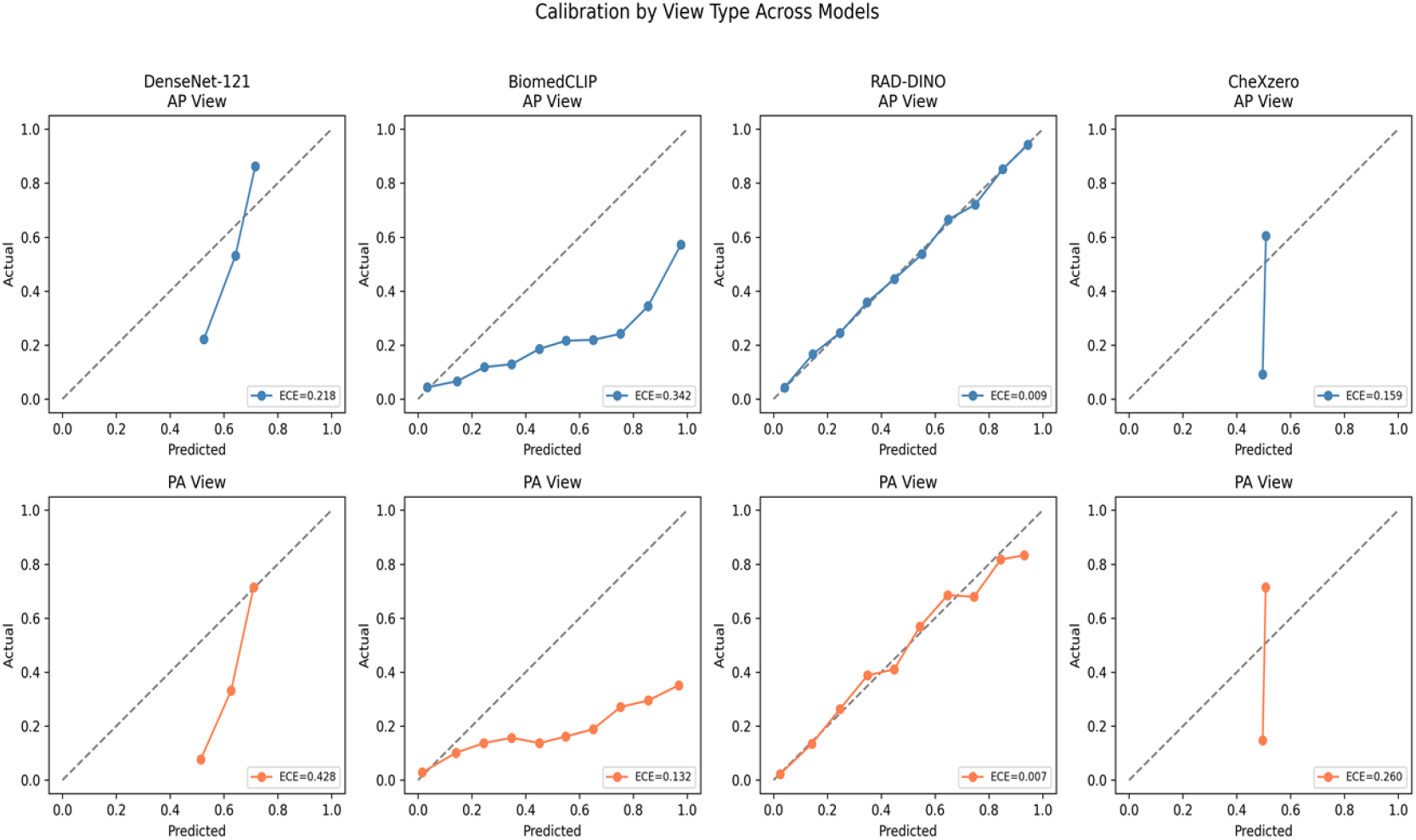
Calibration Curves by View Type Across Models. Reliability diagrams showing predicted probability (x-axis) versus observed frequency (y-axis) for AP views (top row) and PA views (bottom row). Dashed diagonal represents perfect calibration. RAD-DINO achieves near-perfect calibration on RSNA (ECE<0.01), while other models show substantial miscalibration.

### Clinical Impact Estimation

To contextualize these findings, we estimated the clinical impact of view type bias under a hypothetical scenario of 10,000 pneumonia patients with equal AP/PA distribution (Table 5). On RSNA data, BiomedCLIP would result in 1,805 excess missed diagnoses on PA views compared to a hypothetical model with equal sensitivity. On NIH data with its larger view type gaps, the clinical impact was even more pronounced, with BiomedCLIP resulting in 2,475 excess missed diagnoses on PA views.

**Table 5.** Estimated Clinical Impact per 10,000 Pneumonia Patients.

| Model | PA Detected | PA Missed | Excess PA Missed | OR |
| --- | --- | --- | --- | --- |
| **RSNA Dataset** |  |  |  |  |
| CheXzero | 3,770 | 1,230 | 715 | 2.84 |
| RAD-DINO | 3,285 | 1,715 | 1,260 | 5.21 |
| DenseNet-121 | 2,245 | 2,755 | 1,785 | 5.10 |
| BiomedCLIP | 2,710 | 2,290 | 1,805 | 7.87 |
| **NIH Dataset** |  |  |  |  |
| RAD-DINO | 2,560 | 2,440 | 1,115 | 2.65 |
| DenseNet-121 | 1,680 | 3,320 | 1,945 | 5.26 |
| CheXzero | 1,495 | 3,505 | 2,445 | 8.85 |
| BiomedCLIP | 1,525 | 3,475 | 2,475 | 9.29 |

*Assumes 50% AP / 50% PA distribution. Excess PA Missed = PA Missed - AP Missed. OR = odds ratio for missed diagnosis on PA vs AP view.*

## Discussion

This study provides the first systematic examination of foundation model robustness to technical acquisition parameters in medical imaging, with external validation demonstrating critical limitations of domain-specific training approaches. Our findings challenge the assumption that foundation models eliminate or reduce technical biases inherent in traditional supervised learning approaches.

### Domain-Specific Training: Advantages and Limitations

Our initial RSNA findings suggested that domain-specific training (CheXzero) provided superior robustness to view type variation, with a 2.5-fold smaller gap compared to general biomedical models. This advantage emerged despite a nearly 40-fold difference in training data volume (CheXzero: 377K vs BiomedCLIP: 15M), suggesting that data specificity outweighs data volume for acquisition parameter robustness. However, external validation on NIH ChestX-ray14 revealed a critical limitation: **this advantage does not generalize across datasets with different labeling schemes**.

CheXzero’s dramatic performance deterioration (+34.6 percentage point gap increase) likely reflects overfitting to MIMIC-CXR-specific labeling conventions. MIMIC-CXR reports use similar annotation practices to RSNA, whereas NIH labels derive from NLP extraction of historical reports with different terminology. Additionally, there is a potential semantic mismatch between CheXzero’s text prompts (trained on MIMIC-CXR report language) and NIH’s NLP-extracted labels. The prompts optimized for “pneumonia” or “consolidation” may not align well with NIH’s “Infiltration” label, which encompasses a broader radiological finding. This suggests that CheXzero learned to exploit dataset-specific correlations rather than fundamental radiological patterns.

### Self-Supervised Learning as the Path to Generalizable Robustness

RAD-DINO emerged as the most generalizable architecture, demonstrating consistent view type gaps across both datasets (25.2% → 22.3%, −2.9% change). This stability likely reflects DINOv2 [6]’s self-supervised training paradigm, which learns visual representations without text supervision. By avoiding text-based learning signals, RAD-DINO may learn more fundamental visual features that generalize across labeling schemes. RAD-DINO also demonstrated excellent calibration stability (ECE difference <0.01 between views on RSNA, 0.005 on NIH), suggesting that self-supervised training may be preferable for clinical deployment where probability estimates inform decision-making.

### Prompt Engineering: An Accuracy-Fairness Trade-off

Our prompt engineering analysis for BiomedCLIP (Supplementary Table S3) revealed an inherent accuracy-fairness trade-off that may apply to CheXzero as well. While the “Report Style” prompt reduced BiomedCLIP’s view type gap from 35.2% to 16.2%, this came at the cost of substantially reduced AP sensitivity (81.6% vs 90.6% with standard prompts). This demonstrates that while prompts can modulate the gap, they cannot eliminate it without sacrificing overall performance—suggesting the bias is embedded in the model’s learned visual representations, not merely in text-image alignment.

### Implications for Foundation Model Development

These findings challenge assumptions about foundation model development strategies:

1. **Domain specificity is not sufficient:** Training on domain-specific data improves performance within that domain but may reduce external generalizability.
2. **Text supervision introduces label-dependence:** Vision-language models learn correlations between images and text labels, making them sensitive to labeling conventions.
3. **Self-supervised learning may be more robust:** By learning from images alone, self-supervised models avoid text-based shortcuts.
4. **External validation is essential:** Single-dataset evaluations may overestimate model robustness, particularly for domain-specific architectures.

### Regulatory and Clinical Implications

Our finding that 31% of PA pneumonia cases were missed by all four models on external data has direct clinical implications. Current FDA guidance emphasizes demographic subgroup analysis [12], but our results suggest that acquisition parameter auditing should be mandatory for chest X-ray AI regulatory submissions. Furthermore, the dramatic performance differences between internal and external validation highlight the need for multi-site evaluation requirements. Models showing excellent performance on their training distribution may fail systematically on data from different institutions or labeling practices.

### Limitations

Several limitations merit consideration. First, NIH labels derive from NLP extraction and may differ in accuracy from expert annotations. However, the consistency of view type effects across multiple label definitions suggests this is not a major confounder. Second, we evaluated a limited set of foundation models; emerging architectures may show different robustness patterns. Third, the lower AUC values on NIH compared to RSNA reflect the different labeling schemes rather than absolute model quality.

## Conclusions

Foundation models do not eliminate technical acquisition parameter biases in chest X-ray AI. While domain-specific training (CheXzero) provided superior robustness on internal validation, this advantage **collapsed on external data**. Self-supervised learning (RAD-DINO) demonstrated the most generalizable robustness, with consistent view type gap stability across datasets with different labeling schemes (25.2% → 22.3%, despite substantial AUC differences).

These findings have four key implications: (1) external validation across institutions with different labeling practices is essential for robustness claims; (2) self-supervised training may be preferable to vision-language approaches for generalizable medical imaging AI; (3) acquisition parameter auditing should be incorporated into AI regulatory frameworks alongside demographic subgroup analysis; and (4) **the systematic 31% miss rate on PA pneumonia cases across all models represents a patient safety concern** that warrants urgent attention in clinical AI deployment.

## Data Availability

The data can be accessed at https://github.com/hayden-farquhar/FM-robustness

## Data Availability

The RSNA Pneumonia Detection Challenge dataset is publicly available through Kaggle (https://www.kaggle.com/c/rsna-pneumonia-detection-challenge). The NIH ChestX-ray14 dataset is publicly available from NIH (https://nihcc.app.box.com/v/ChestXray-NIHCC). Analysis code and model predictions is available at https://github.com/hayden-farquhar/FM-robustness.

## Acknowledgments

None.

